# Estimates of excess mortality for the five Nordic countries during the Covid-19 pandemic 2020-2021

**DOI:** 10.1101/2022.05.07.22274789

**Authors:** Kasper P. Kepp, Jonas Björk, Vasilis Kontis, Robbie M. Parks, Kristoffer T. Bæk, Louise Emilsson, Tea Lallukka

## Abstract

Excess mortality during the Covid-19 pandemic are of major scientific and political interest. Here we review all-cause excess mortality estimates from different methods for the five Nordic countries (Denmark, Finland, Iceland, Norway, and Sweden), which have been much studied during the Covid-19 pandemic. In the comparison of the methods, we use simple sensitivity estimates and linear extrapolations of the death data to discuss uncertainties and implications for reporting ratios and infection fatality rates. We show using back-calculation of expected deaths from Nordic all-cause deaths that a recent study in *Lancet*, which is a clear outlier in the overviewed estimates, most likely substantially overestimates excess mortality of Finland and Denmark, and probably Sweden. The other estimates are more consistent and suggest a range of total Nordic excess deaths of perhaps 15−20,000, a more uniform ability to identify Covid-19-related deaths, and more similar infection fatality rates for the Nordic countries. All these estimates should be taken with caution in their interpretation as they miss detailed account of demographics, such as changes in the age group populations over the study period.

## Introduction

Excess mortality (the difference between observed and expected number of deaths) during the Covid-19 pandemic are of major scientific and political interest, as they provide objective estimates of the pandemic burden not affected by different testing procedures and and varying registration criteria of Covid-deaths between and within countries.^1–4^ All-cause excess mortality includes deaths due to Covid-19 after infection by severe acute respiratory syndrome coronavirus 2 (SARS-CoV-2) and other causes such as cancer^5^ and is of interest for a total evaluation of pandemic impact. Because of their major importance, detailed and critical review of methods to obtain excess mortality, the numbers, and their implications, should be of high priority. Here, we provide such a systematic review of several methods to estimate the all-cause excess mortality for 2020 and 2021 for the five Nordic countries (Denmark, Finland, Iceland, Norway, and Sweden).

We studied the Nordic countries due to i) special interest and insight by the authors, ii) they form a historically and culturally related entity with public health data of high quality, iii) they have been much studied during the pandemic, with claims of both failures (Sweden) and successes (e.g., Norway and Denmark)^6–8^, and iv) all five countries have final annual all-cause deaths available for 2020 and 2021.

Our review is partly motivated by a recent paper in Lancet by Wang et al.^9^ using a model for predicting global excess mortality, which concluded that excess mortalities of Sweden, Denmark, and Finland were much larger^9^ than previously estimated,^8,10^ with excess deaths per capita of Denmark (and almost Finland) similar to Sweden, and very large differences in the five countries’ ability to identify Covid-19 deaths, with ratios of excess deaths to official Covid-19 deaths of 3.2 and 5.0 for Denmark and Finland but -8.5 and 0.6 for Iceland and Norway.^9^ While registration criteria are never perfect,^11^ these differences are surprisingly large and invite further analysis. Another surprising consequence, when combined with infection estimates, is 6−7 times higher infection fatality rates (IFR) in Finland and Denmark than in Norway, and almost double that of Sweden.^12^ Due to the topic’s importance, these major differences warrant scrutiny.

To better understand the different model results, we use the latest official administrative register data to examine the death estimates via linear extrapolation. Specifically, we use annual all-cause Nordic death data to back-calculate the expected deaths required (but not reported) for stated excess deaths to be accurate. Second, we review the different model estimates and discuss limitations and realistic ranges of the events.

## Methods

### Data used

We collected the final all-cause deaths for 2010−2021 from the relevant statistics authorities, divided into years in order to avoid seasonal effects (we note that Wang et al.^9^ used both weekly and monthly data and they did not have access to the final 2021 data in their estimates), **Table S1**, as well as mean population data per year. The links to the sources of the data can be found in the Data availability statement.

### Back-calculating expected deaths

Excess deaths are defined as observed real deaths subtracted by the expected deaths, equation (1):

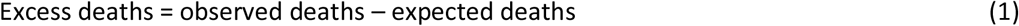

To test the validity of the model numbers we used reported excess deaths and the actual, final deaths for back-calculating the expected deaths implied via equation (2):

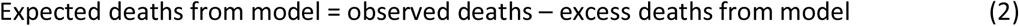

We then compared these implied expected deaths with the actual death data to test the reasonability of assumed baselines of estimated deaths, and conversely, the stated excess deaths.

### Challenges estimating excess mortality

Challenges when estimating excess mortality include: 1) the trends in population structure, notably changes in aging and demographic shifts, should be accounted for; 2) depending on time period, seasonality and week/year overlap (ISO-week) if using weekly data vs. annual data (mostly a minor effect); 3) mortality displacement, with mortality in one time period correlating with the next period^13,14^; 4) unusual recent events could distort baseline estimates. If interested in individual causes of death, additional assumptions emerge as true causes of death in the elderly are frequently multi-factorial and difficult to establish, and such assessment is beyond the scope of this study.

No method handles all issues perfectly. Linear extrapolation on full-year data solves some of the issues as it averages out season and can handle population structure on short timescales but is sensitive to recent unusual events, as analyzed below. Yet this approach, as applied e.g. by Karlinsky and Kobak,^10^ and The Economist,^15^ does not include any assumptions beyond linearity and mortality displacement is reasonably accounted for by linear regression compromising low and high death years. They are thus important as sensitivity tests for understanding and comparing the data, which is essential.^16^ Methods that use fixed functional (e.g., sinusoidal) forms to estimate the baseline and reduce the impact of unusual influenza seasons or heat waves also exist, e.g., Euromomo.^17,18^

### Sensitivity analysis and comparisons

To understand ranges and uncertainties, we calculated 5-year and 10-year linear trends in all-cause deaths (2015−2019 and 2010−2019). The removal of single unusual years provides an estimate of maximum baseline impact and was done for the recent years 2018 and 2019.

Changes of the population age have a large impact on expected deaths (and thus deduced excess deaths)^19^, with death rates being exponential in age, also for deaths due to covid-19,^20^ and can be accounted for using mortality rates based on mean annual age-group-specific populations from the Nordic Council’s aggregate data from the five statistical departments (see https://pxweb.nordicstatistics.org/). However, all estimates reviewed here only report total all-cause mortality, so comparison of age-specific mortality was not possible.

Excess mortality estimates for 2020 and 2021 were compiled from the method of Wang et al.^9^ This model estimates expected deaths via a 6-model ensemble that tries to correct for missing data due to late registration and leaves out heat waves of three weeks inside the time series. It also applies a global statistical model to countries that do not have data available. Since this model is affiliated with the Institute for Health Metrics and Evaluation it is referred to as IHME below. For more details on this method, we refer to the Appendix of Wang et al.^9^

We additionally studied the World Mortality Dataset (WMD; Ariel Karlinsky and Dmitry Kobak)^10^ whose results may partly differ despite the similarities in the design of the models (see below). These models run weekly from December 30, 2019 to January 2, 2022, giving four days difference in total deaths relative to the yearly time series in **Table S1**, and use linear trends to estimate expected deaths.

We also reviewed estimates from the method used by The Economist (via Sondre Solstad) in two different versions;^15^ one that includes the January and February 2020 death data in fitting the expected death trends, and one that does not. The machine learning (gradient boosting) model instead uses hard data when available as for the Nordics, based on the direct (not log-linear) deaths, and using expected deaths in a similar way as the WMD approach.^10^

We also included the estimates of the 2020 and 2021 excess mortality by the World Health Organization (WHO) Technical Advisory Group for COVID-19 Mortality Assessment, released May 5, 2022.^21^ These estimates are based on a statistical Poisson-type model that, as WMD and Economist models, emphasizes the hard data for countries where these are available, which includes the Nordics, and predictions for those where they unavailable.^22^ The model in the first cases uses loglinear fitting with time variation modelled via splines. More details on these models are available elsewhere.^22^

In addition, we included estimates from an ensemble of Bayesian methods,^23,24^ referred to below as “Bayesian model ensemble” (BME). This method uses weekly deaths and populations from Eurostat and an ensemble of Bayesian probabilistic models to estimate the expected number of deaths in the absence of the pandemic.^23^ The models were designed to account for medium-long-term secular trends in mortality, the potential dependency of death rates in each week on those in preceding week(s) and in each year on those in preceding year(s), and factors that affect mortality, including seasonality, temperature, and public holidays. The models were fitted to up-to-date data from 2010 until the last week of December 2019 and then used to generate predictions of expected deaths in 2020-2021.

We did not include Euromomo^17^, as its all-cause excess mortalities for 2020 and 2021 are not available. Since Euromomo removes Winter periods when fitting expected mortality, its baselines are lower than all-cause baselines and the model tends to produce excess mortality every year. Thus, one needs to back-add influenza deaths or fit to the full data to appropriately compare to all-cause excess death estimates.

## Results and discussion

### Overview of mortality estimates

**Table 1** shows an overview of the analyzed data for the five Nordic countries. While most methods are in some relative agreement, one of the methods reviewed, the new study by Wang et al.^9^ in Lancet, produces very different results from other models studied, with potential major implications, perhaps best seen from the excess deaths per 100,000 people in **Table 2**, with IHME estimates for Denmark, Finland, and Sweden being much higher and similar than for other methods.

**Table 1.**
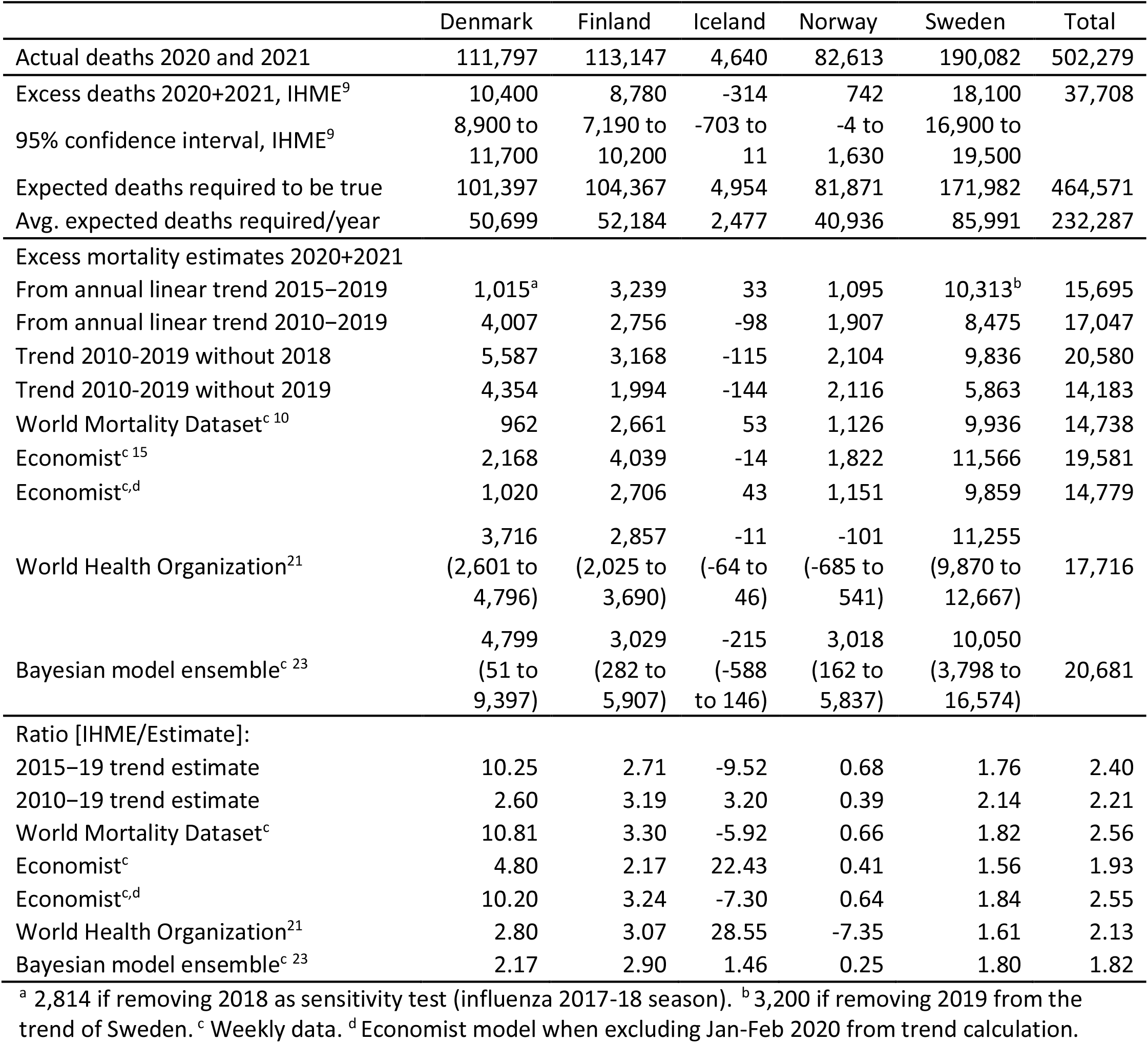
Summary of data for the five Nordic countries.

|  | Denmark | Finland | Iceland | Norway | Sweden | Total |
| --- | --- | --- | --- | --- | --- | --- |
| Actual deaths 2020 and 2021 | 111,797 | 113,147 | 4,640 | 82,613 | 190,082 | 502,279 |
| Excess deaths 2020+2021, IHME<sup>9</sup> | 10,400 | 8,780 | -314 | 742 | 18,100 | 37,708 |
| 95% confidence interval, IHME<sup>9</sup> | 8,900 to<br>11,700 | 7,190 to<br>10,200 | -703 to<br>11 | -4 to<br>1,630 | 16,900 to<br>19,500 |  |
| Expected deaths required to be true | 101,397 | 104,367 | 4,954 | 81,871 | 171,982 | 464,571 |
| Avg. expected deaths required/year | 50,699 | 52,184 | 2,477 | 40,936 | 85,991 | 232,287 |
| Excess mortality estimates 2020+2021 |  |  |  |  |  |  |
| From annual linear trend 2015–2019 | 1,015<sup>a</sup> | 3,239 | 33 | 1,095 | 10,313<sup>b</sup> | 15,695 |
| From annual linear trend 2010–2019 | 4,007 | 2,756 | -98 | 1,907 | 8,475 | 17,047 |
| Trend 2010-2019 without 2018 | 5,587 | 3,168 | -115 | 2,104 | 9,836 | 20,580 |
| Trend 2010-2019 without 2019 | 4,354 | 1,994 | -144 | 2,116 | 5,863 | 14,183 |
| World Mortality Dataset<sup>c 10</sup> | 962 | 2,661 | 53 | 1,126 | 9,936 | 14,738 |
| Economist<sup>c 15</sup> | 2,168 | 4,039 | -14 | 1,822 | 11,566 | 19,581 |
| Economist<sup>c,d</sup> | 1,020 | 2,706 | 43 | 1,151 | 9,859 | 14,779 |
| World Health Organization<sup>21</sup> | 3,716<br>(2,601 to<br>4,796) | 2,857<br>(2,025 to<br>3,690) | -11<br>(-64 to<br>46) | -101<br>(-685 to<br>541) | 11,255<br>(9,870 to<br>12,667) | 17,716 |
| Bayesian model ensemble<sup>c 23</sup> | 4,799<br>(51 to<br>9,397) | 3,029<br>(282 to<br>5,907) | -215<br>(-588<br>to 146) | 3,018<br>(162 to<br>5,837) | 10,050<br>(3,798 to<br>16,574) | 20,681 |
| Ratio [IHME/Estimate]: |  |  |  |  |  |  |
| 2015–19 trend estimate | 10.25 | 2.71 | -9.52 | 0.68 | 1.76 | 2.40 |
| 2010–19 trend estimate | 2.60 | 3.19 | 3.20 | 0.39 | 2.14 | 2.21 |
| World Mortality Dataset<sup>c</sup> | 10.81 | 3.30 | -5.92 | 0.66 | 1.82 | 2.56 |
| Economist<sup>c</sup> | 4.80 | 2.17 | 22.43 | 0.41 | 1.56 | 1.93 |
| Economist<sup>c,d</sup> | 10.20 | 3.24 | -7.30 | 0.64 | 1.84 | 2.55 |
| World Health Organization<sup>21</sup> | 2.80 | 3.07 | 28.55 | -7.35 | 1.61 | 2.13 |
| Bayesian model ensemble<sup>c 23</sup> | 2.17 | 2.90 | 1.46 | 0.25 | 1.80 | 1.82 |

<sup>a</sup> 2,814 if removing 2018 as sensitivity test (influenza 2017-18 season). <sup>b</sup> 3,200 if removing 2019 from the trend of Sweden. <sup>c</sup> Weekly data. <sup>d</sup> Economist model when excluding Jan-Feb 2020 from trend calculation.

**Table 2.** Excess mortality estimates 2020+21 per 100,000 people (using population January 1, 2021).

|  | Denmark | Finland | Iceland | Norway | Sweden |
| --- | --- | --- | --- | --- | --- |
| Excess deaths per 100,000 | 5,840,045 | 5,533,793 | 368,792 | 5,391,369 | 10,379,295 |
| IHME<sup>9</sup> | 178 | 159 | -85 | 14 | 174 |
| Trend 2015–2019 | 17 | 59 | 9 | 20 | 99 |
| Trend 2010–2019 | 69 | 50 | -27 | 35 | 82 |
| Trend 2010–2019 without 2018 | 96 | 57 | -31 | 39 | 95 |
| Trend 2010–2019 without 2019 | 75 | 36 | -39 | 39 | 56 |
| World Mortality Dataset<sup>10</sup> | 16 | 48 | 14 | 21 | 96 |
| Economist<sup>15</sup> | 37 | 73 | -4 | 34 | 111 |
| Economist<sup>a</sup> | 17 | 49 | 12 | 21 | 95 |
| World Health Organization<sup>21</sup> | 64 | 52 | -3 | -2 | 108 |
| Bayesian model ensemble<sup>23</sup> | 82 | 55 | -58 | 56 | 97 |

<sup>a</sup> Economist model excluding Jan-Feb 2020 from trend calculation.

To understand these differences in more detail, we used linear extrapolations to estimate what the expected deaths would have been if they followed a trend in the actual annual death data and compared these to the final Nordic annual deaths for 2020 and 2021 to estimate what the excess mortality would correspondingly be via Equation (2) and subject these extrapolations to sensitivity tests of time-period and unusual years.

**Figure 1** shows the actual all-cause annual deaths of the five Nordic countries for the years 2010-2021, updated as of April 27, 2022. We added a red line for each country indicating the average expected deaths of 2020 and 2021 required for the excess mortality estimated by Wang et al.^9^ to be true, using Equation (2). As seen from **Figure 1**, the implied expected deaths (red lines) seem inconsistent with the actual data for the years prior to 2020 for Denmark, Finland, and Sweden. In all three cases, the expected all-cause deaths are substantially underestimated relative to both 5-year and 10-year trends of the data. For Denmark and Sweden, the implied expected deaths are lower than any observed deaths the previous 10 years despite a recent increasing trend. A similar result is seen for mortality rates that account for changing population size, **Figure S1** (calculated as in **Table S2**). Thus, we conclude that the estimates are unlikely to be realistic.

**Figure 1.**
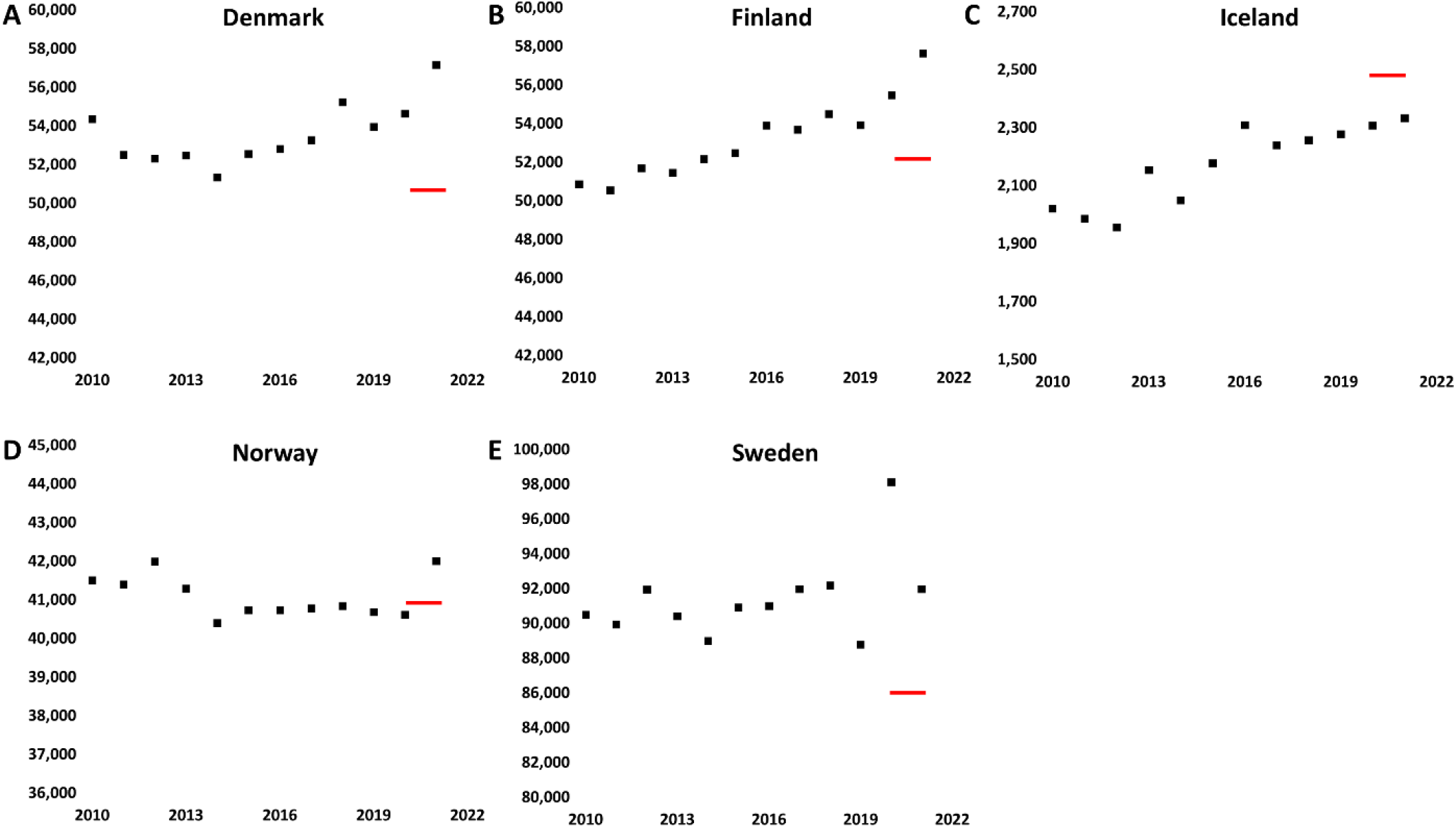
All-cause deaths of the Nordic countries 2010-2021 (squares). (**A**) Denmark. (**B**) Finland. (**C**) Iceland. (**D**) Norway. (**E**) Sweden. The red lines show the back-calculated expected deaths (average of 2020 and 2021) implied by the excess deaths in Wang et al.^9^, using Equation (2) (analysis for mortality rates in **Figure S1**).

### Estimates of sensitivities

To understand the reviewed excess mortality estimates further, we supplemented them with some sensitivity tests. To do so, we used the annual Nordic all-cause death data to compute simple excess death estimates with 5- or 10-year linear trends as sensitivity estimates of the impact of time-period and tested the sensitivity to leaving out recent years with large potential impact (**Figure S1**).

Two special years are notable: 1) Some countries had a particularly deadly 2017-18 influenza season^25^ that is clearly visible in the raw death data in **Figure 1** of Denmark (Nordic influenza deaths typically cluster in January to March even if the season starts earlier). 2) Sweden had unusually low mortality in 2019, also clearly visible in **Figure 1**. The extrapolations without 2018 or 2019 show relatively little impact on Finland, Iceland, and Norway’s deaths, but large effect for Denmark and Sweden, indicating that the excess deaths of the two latter countries are more difficult to estimate. Methods that do not account for these unusual years may suffer uncertainties as implied in **Table 1**.

In principle, special periods of unusual low or high mortality could be smoothed out, but such removals could also produce errors due to mortality displacement (time correlations of deaths).^14,26^ If one discards 2019 completely, a maximum estimate of the impact of this, Sweden’s excess mortality would be substantially lower. While Sweden experienced less mortality in 2019 and more in 2020, other Nordic countries had lower mortality in 2020, as noted previously,^8,10,23^ but relatively more in 2021 (**Figure 1**). This could suggest mortality displacement^27^ or e.g., immunity effects, although this needs to be explored further.

For Iceland, estimates also differ substantially percentwise partly due to the small numbers involved and to fluctuations, but the IHME estimate is still far from any other estimate in **Table 1. Figure 1** suggests that the implied baseline is rather high. In total, the excess mortality reported by Wang et al.^9^ for the five Nordic countries is more than a factor 2 of that deduced from the 5-year or 10-year trends, and this difference is not reduced by leaving out the most impactful special years.

### Comparison of models

The total excess mortality estimates for 2020 and 2021 from the World Mortality dataset (WMD)^10^ were compiled as in **Table 1**. This method uses linear extrapolations and thus carries the types of uncertainties analyzed above, in **Table 1**. The WMD estimates agree well with the annual data trends as expected due to their similar methodology, with variations far from the estimates by Wang et al. In total, numbers of the IHME model are 2.5-fold that of the WMD, which is an enormous difference if both models are fitted to similar available data for the Nordic countries (**Table 1**).

We also reviewed the estimates of two Economist models. Wang et al. provide a double-logarithmic plot of absolute excess deaths (their Figure S5) to suggest agreement between their data and the Economist, but such a plot is dominated by large countries, making discrepancies for individual countries less clear. **Table 1** lists the Economist estimates both with and without the first two months of 2020 included when estimating baselines, which has a notable, relevant impact. Still, these estimates are far from those of IHME: For example, the Economist estimate for Denmark is less than a fourth of the 10,400 suggested by IHME.

On May 5, 2022, the WHO updated their detailed estimates of excess mortality for 2020 and 2021;^21^ these data in **Tables 1/2** also show good agreement with the ranges of other methods, except having a remarkable, somewhat lower excess mortality for Norway. We find that this method also gives total excess mortality for the five Nordic countries combined of approximately half that of IHME.^9^ Despite the variations in **Table 1**, the IHME estimates are outside the ranges of all other methods for Sweden, Finland, and Denmark. For example, for Denmark, the IHME estimate is 8,900−11,700, i.e., even the smallest number is much larger than other ranges in **Table 1**. In this light, the narrow IHME confidence intervals far from other ranges are concerning. Although Wang et al. did not separate years, their excess deaths also seem high vs. other estimates for earlier parts of the pandemic listing a few thousand excess deaths for Denmark and Finland.^28^

The Bayesian model ensemble^23,24^ also has its central estimates relatively similar to those of the other methods, with the exception that it gives the highest central estimate for Norway, although the confidence interval spans from 162 to 5,837, i.e., still within the range of all other methods (**Table 1**). For the other countries, the Bayesian model ensemble is in relatively good agreement with the other methods except IHME, and the sum of the median estimates, 20,681, is in reasonable agreement with these other models. We also note that the lower IHME estimates for Finland and Sweden are well above the upper end of the 95% uncertainty interval for both WHO and the Bayesian model ensemble. The average total for the two Economist models, WMD, WHO, and BME is 17,499 with a standard deviation of 2717. The 95% confidence interval for all nine non-IHME estimates is 17,222 ± 1974 (15,248−19,197).

### The Nordic countries’ capacity to identify Covid-19 deaths

Wang et al.^9^ suggested that Nordic countries had enormous differences in their ability to identify deaths due to Covid-19, with a ratio between excess and official deaths of 3.2 and 5.0 for Denmark and Finland, but only 0.6 for Norway and 1.2 for Sweden^9^. While we expect differences due to different reporting strategies, knowing the Nordic healthcare systems and pandemic responses, the many-fold under-registration seems implausible to us, as does the major heterogeneity in this capacity, although this needs to be explored further in future work.

We calculated this ratio as shown in **Figure 2** (raw ratios are summarized in **Table S3**), using the official deaths until December 31, 2021. We find that the Nordic countries’ ability to identify Covid-19 deaths (assuming most excess deaths are Covid-19 and that the assumptions of the models are fulfilled) is much more homogeneous with the other estimates than with the IHME model.^9^ It is the only model that estimates that Sweden had more excess deaths than official Covid-19 deaths, and in particular the apparent ability of Finland and Denmark to identify their Covid-19 deaths is much more similar to other countries for the other studied estimates, whereas the ratios for Iceland are highly fluctuating and uncertain, due to the relatively large spread in absolute estimates relative to the overall small numbers markedly affecting the ratio.

**Figure 2.**
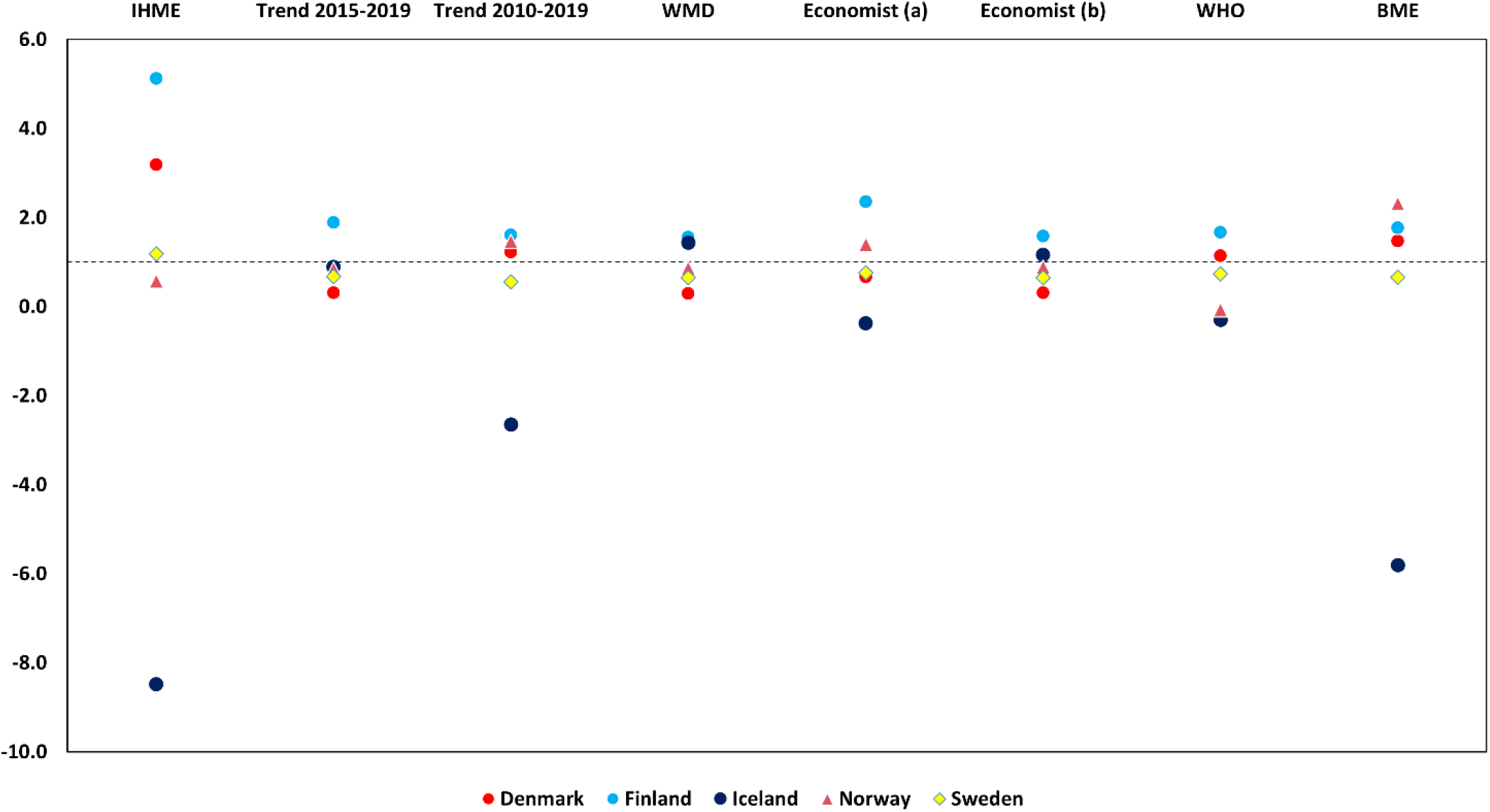
Estimated excess deaths divided by official Covid-19 deaths for 2020 and 2021. Economist (a) and (b) refer to the model with and without Jan−Feb 2020 included when estimating expected deaths. The dotted line shows the hypothetical ratio 1 (identical official and excess deaths).

### Impact of population structure

The Nordic countries differ somewhat in age structures, as shown in the mean population of 1-year age groups in 2020 in **Figure S2**. The fraction of people >70 years varied from 9.9% in Iceland to 16.1% in Finland in 2020 when the pandemic started (**Table S4**). **Figure 3** shows the death rates of the 5-year groups based on total deaths and the mean population of each age group (numbers in **Table S5**, log-plot in **Figure S4** for relative changes). By far most excess mortality is observed in the 70+ age groups, consistent with the exponential impact of age on (Covid-19) mortality^20^. Different changes of the populations of the age groups from 2010-2019 (**Figure S3**) are thus important for understanding expected deaths, rationalizing excess deaths, and performance comparisons or policy implications. As such analysis is not done by any of the reviewed models, the numbers summarized in this work should be interpreted with caution given the possible impact of such variations in demographic development.

**Figure 3.**
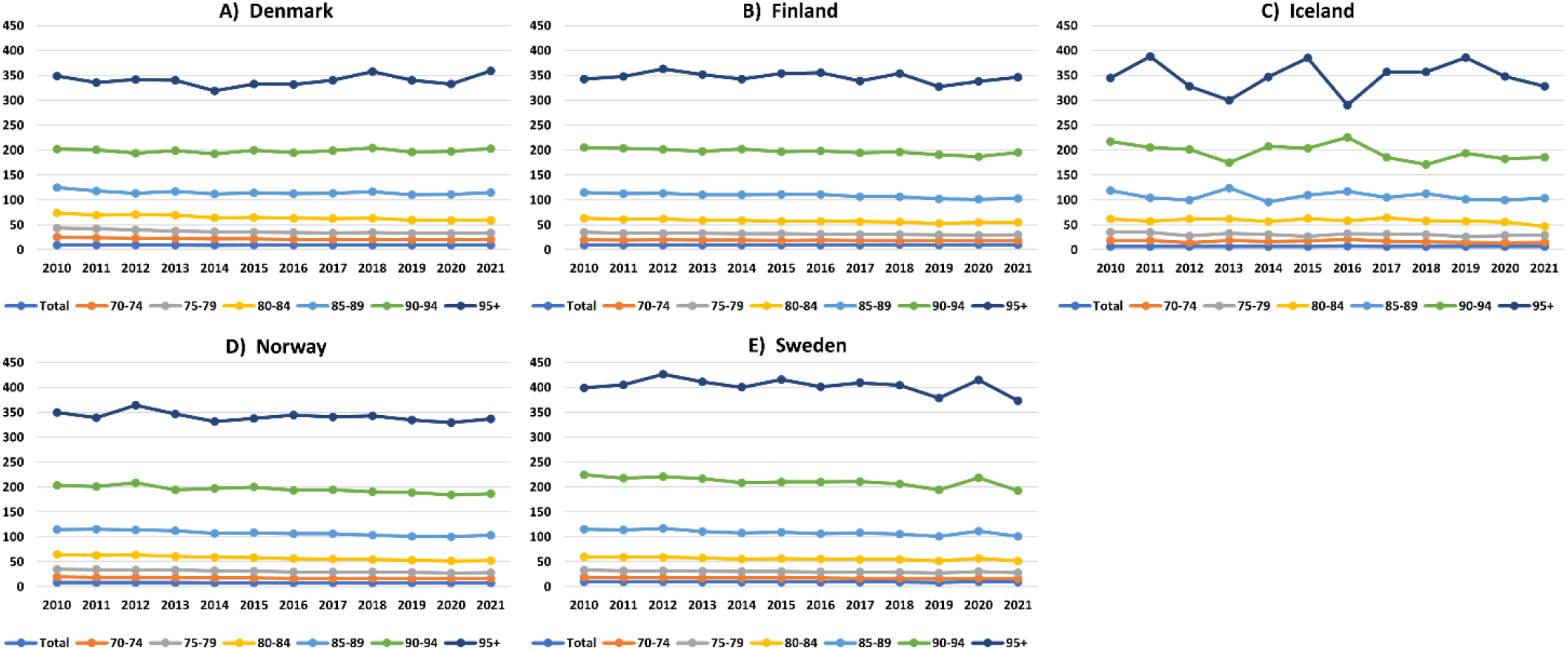
Nordic age-specific mortality rates per 1000 people within five-year age groups for 70+ years. Based on final official deaths until 2021 per age group and mean annual population of each age group. (**A**) Denmark. (**B**) Finland. **(C)** Iceland. **(D)** Norway. **(E)** Sweden. (Logarithmic plot in **Figure S4**).

### Implications for infection fatality rates

Death estimates also have consequences for IFR values (the fraction of total infections in the country that leads to death) to the extent they are mainly due to Covid-19. A recent paper estimated global infection until November 14, 2021, before the Omicron variant (Barber et al.),^12^ with infection estimates partly based on e.g., seroprevalence, that seem reasonable compared to the historic Nordic seroprevalence estimates, with Sweden having approximately double the infection of the other Nordic countries up to November 2021 (22.4%, vs. 7.9-13.5% **Table S6**).

However, when IHME-modelled deaths (for the shorter period, and attempted to be corrected for non-Covid-19 cases) were applied to these infection estimates, surprising IFR values resulted, with e.g., average infections in Denmark and Finland being 6-7 times more deadly than in Norway and almost doubly as deadly as in Sweden (**Table S6**).^12^ Such differences are hard to explain by population or healthcare variations, pandemic management or vaccination strategies of the Nordic countries, and warrant further scrutiny.

To very roughly estimate what the corresponding IFR values would have been for the other methods reviewed here, we used the IFR estimated by Barber et al. that used the IHME model attempted corrected for non-Covid-19 cases, and our scale factors calculated in the bottom of **Table 1**, under the simple assumption that the other estimates follow the same scaling and non-Covid-19 corrections for the time period, i.e., that most excess deaths are due to Covid-19 also for late 2021 and that the fraction is mostly independent of model.

**Figure S5** shows these values as a crude indication of the IFR that would be implied by other estimates and models. The average IFR estimates for all other methods are 0.29 ± 0.19% for Denmark, 0.52 ± 0.08% for Finland, 0.01 ± 0.03% for Iceland, 0.39 ± 0.26% for Norway, and 0.39 ± 0.04% for Sweden, compared to the IFR values from IHME, varying up to 1.2%. In other words, the other methods imply more similar IFR values, if one assumes the ratio of excess to true covid-19 cases from Barber et al. applies to these models also (in the limit that it is instead a fixed number of non-Covid deaths rather than a proportion, the discrepancy would be even larger). The IFR estimates by O’Driscoll et al.^20^ for the first part of 2020 ranged from perhaps 0.5-0.7% for Denmark, Norway, Finland, and Sweden, largely consistent with these crude estimates, at least regarding the more homogenous IFR; more lethal variants, alpha and delta during 2021, but also mass vaccination, have changed these IFR values, but it is hard to reconcile the similar IFR estimates for 2020 from O’Driscoll et al.^20^ with the cumulative IFRs until late 2021 from Barber et al.^12^

In summary, combining **Figure 2** and **Figure S5**, we find it particularly anomalous that the same two countries, Finland and Denmark, that would be worse at identifying their Covid-19 deaths by factors of at least 2-4 vs. e.g. Sweden, or 4-8 vs. Norway (**Figure 2**) would also simultaneously be the two countries where every SARS-CoV-2 infection had perhaps twice the lethality of infections in Sweden and 6-7 times higher fatality than infections in Norway (**Figure S5**), which also seems inconsistent with O’Driscoll et al. even considering the different time period and age-stratification. The parsimonious explanation to these anomalies is that the IHME excess mortality estimates may not be accurate for these countries, even beyond the implications of **Figure 1**. Regardless, the enormous heterogeneity in these estimates is problematic given their importance, and we invite further analysis of the methods to address this heterogeneity.

## Conclusions

Given the scientific and political importance of excess mortalities, we consider systematic and open critical comparison of different methods to estimate these of very high priority. Our paper constitutes the first such effort to our knowledge, which we hope will be followed by others. We note that the total excess numbers as reviewed here cannot directly inform performance estimates or policy implications even if they were accurate, as they miss context on demographics such as e.g., population age changes over time.

We reviewed estimates of the excess mortality during the pandemic 2020 and 2021 for the Nordic countries Denmark, Finland, Iceland, Norway, and Sweden, which have been of much interest as both possible successes and failures, as an ideal study case due to their high-quality data and similarities. Our purpose was not to provide new advanced estimates, but to critically review methods and estimate uncertainties, limitations, and also implications of the numbers, especially due to recent debate on per capita deaths and registration differences, and our study should only be seen in this specific context.

As one of the methods (IHME, Wang et al.^9^) produces very distinct results from all other studied estimates, additional analysis of this estimate was done. By back-calculating expected deaths we show that IHME numbers^9^ seem inconsistent with actual data. Accordingly, excess mortalities seem substantially overestimated relative to reasonable variations in the data for Finland and Denmark in particular, and also for Sweden. We find that the main uncertainties in determining the excess deaths are the 2018 influenza, especially for Denmark and a bit less Finland, and the low Swedish 2019 mortality.

Our review of methods and sensitivity tests suggest that the overall excess mortality in the Nordic countries like range between 15,000 and 20,000. The WHO estimates that came out just on the last days of this paper’s production (May 5, 2022) are in the middle of this range (17,716)^21^. The Bayesian model ensemble^23^ gives a result of 20,681, and a variety of linear regressions produce relatively similar results. These numbers are approximately half that suggested by the IHME model and imply much more similar capability of identifying Covid-19 deaths and more homogeneous IFR values, which we believe more consistent with expectations of pandemic management, e.g., the relevance of postponing infection until vaccines were more widely available in the Nordic countries.

The heterogeneity in estimates even for countries with very good data as reviewed here invites concern over estimates for countries without good data and documents the importance of careful and public evaluation and comparison of methods to calculate excess mortality and a resolute need for data and method transparency, in particular metrics from training and external validation, and critical discussion of results. We hope that our study will set such a precedence for the future.

More generally, our study illustrates the need for data-focused quality control of complex models whose uncertainties and assumptions may be difficult to interpret. For policy implications and for wider public it is important to have a clear messaging, but high-quality data should not be subordinate to complex models. We warmly invite further studies that account in more detail for these topics and uncertainties.

## Data Availability

All data required for the calculations in this work are available at the web pages of Statistics Denmark, Statistics Norway, Statistics Sweden, Statistics Finland, and Statistics Iceland, and respective method data pages.
Statistics Finland: https://pxweb2.stat.fi/PxWeb/pxweb/en/StatFin/StatFin__kuol/statfin_kuol_pxt_12ak.px/
Statistics Iceland: https://px.hagstofa.is/pxen/pxweb/en/Ibuar/Ibuar__Faeddirdanir__danir__danir/MAN05210.px/table/tableViewLayout1/?rxid=247e4620-6490-4f04-b60a-58a68a3afbd9
Statistics Denmark: https://www.statistikbanken.dk/20014
Statistics Norway: https://www.ssb.no/en/statbank/table/08425/
Statistics Sweden: https://www.statistikdatabasen.scb.se/pxweb/en/ssd/START__BE__BE0101__BE0101G/ManadFoddDod/
World Mortality Dataset: https://github.com/dkobak/excess-mortality/blob/main/excess-mortality-timeseries.csv
Economist estimates: https://www.economist.com/graphic-detail/coronavirus-excess-deaths-estimates
WHO estimates: https://www.who.int/data/sets/global-excess-deaths-associated-with-covid-19-modelled-estimates
Comparative Nordic data: mean population sizes, death rates: https://pxweb.nordicstatistics.org/pxweb/en/Nordic%20Statistics/Nordic%20Statistics__Demography__Population%20change/

https://pxweb.nordicstatistics.org/pxweb/en/Nordic%20Statistics/Nordic%20Statistics__Demography__Population%20change/

## Acknowledgements

The authors thank Tauno Tyllinen from Statistics Finland, Tomas Johansson from Statistics Sweden, Bergný Tryggvadóttir from Statistics Iceland, and Dorthe Larsen from Statistics Denmark for communication regarding country data, Sondre Solstad for discussions on the Economist model and the estimates in Table 1, Charles Tallack (The Health Foundation), Jens Nielsen (Statens Seruminstitut), James Wood and David Muscatello (School of Population Health, UNSW) and Theis Lange and Terese Jørgensen (Copenhagen University) for stimulating discussions and feedback, and Dmitry Kobak (Tübingen University) and Ariel Karlinsky (Hebrew University), helpful discussion and confirmation of the WMD data.

## Data availability statement

All data required for the calculations in this work are available at the web pages of Statistics Denmark, Statistics Norway, Statistics Sweden, Statistics Finland, and Statistics Iceland, and the method estimates are available at public sites, as summarized below:

Statistics Finland: https://pxweb2.stat.fi/PxWeb/pxweb/en/StatFin/StatFin__kuol/statfin_kuol_pxt_12ak.px/

Statistics Iceland: https://px.hagstofa.is/pxen/pxweb/en/Ibuar/Ibuar__Faeddirdanir__danir__danir/MAN05210.px/table/tableViewLayout1/?rxid=247e4620-6490-4f04-b60a-58a68a3afbd9

Statistics Denmark: https://www.statistikbanken.dk/20014

Statistics Norway: https://www.ssb.no/en/statbank/table/08425/

Statistics Sweden: https://www.statistikdatabasen.scb.se/pxweb/en/ssd/START__BE__BE0101__BE0101G/ManadFoddDod/

Comparative Nordic data: mean population sizes, mortality rates: https://pxweb.nordicstatistics.org/pxweb/en/Nordic%20Statistics/Nordic%20Statistics__Demography__Population%20change/

World Mortality Dataset: https://github.com/dkobak/excess-mortality/blob/main/excess-mortality-timeseries.csv

Economist estimates: https://www.economist.com/graphic-detail/coronavirus-excess-deaths-estimates

WHO estimates: https://www.who.int/data/sets/global-excess-deaths-associated-with-covid-19-modelled-estimates

Data used for the Bayesian model ensemble: https://github.com/vkontis/excess_mortality/tree/pub2 Weekly deaths and population data (Eurostat): https://ec.europa.eu/eurostat/data/database (tables demo_r_mwk_05 and demo_pjangroup) Temperature (ERA5) and gridded population: https://www.ecmwf.int/en/forecasts/datasets/reanalysis-datasets/era5 https://sedac.ciesin.columbia.edu/data/collection/gpw-v4

## SUPPLEMENTARY INFORMATION

**Table S1.** All-cause annual deaths from the Statistical departments of the Nordic countries 2010−2021.^**a**^.

|  | Denmark | Finland | Iceland | Norway | Sweden |
| --- | --- | --- | --- | --- | --- |
| **2010** | 54,368 | 50,887 | 2,020 | 41,499 | 90,487 |
| **2011** | 52,516 | 50,585 | 1,986 | 41,393 | 89,938 |
| **2012** | 52,325 | 51,707 | 1,955 | 41,992 | 91,938 |
| **2013** | 52,471 | 51,472 | 2,154 | 41,282 | 90,402 |
| **2014** | 51,340 | 52,186 | 2,049 | 40,394 | 88,976 |
| **2015** | 52,555 | 52,492 | 2,178 | 40,727 | 90,907 |
| **2016** | 52,824 | 53,923 | 2,309 | 40,726 | 90,982 |
| **2017** | 53,261 | 53,722 | 2,240 | 40,774 | 91,972 |
| **2018** | 55,232 | 54,527 | 2,257 | 40,840 | 92,185 |
| **2019** | 53,958 | 53,949 | 2,277 | 40,684 | 88,766 |
| **2020** | 54,645 | 55,488 | 2,307 | 40,611 | 98,124 |
| **2021** | 57,152 | 57,659 | 2,333 | 42,002 | 91,958 |

<sup>a</sup> Collected from the web pages in the data availability statement.

**Table S2.**
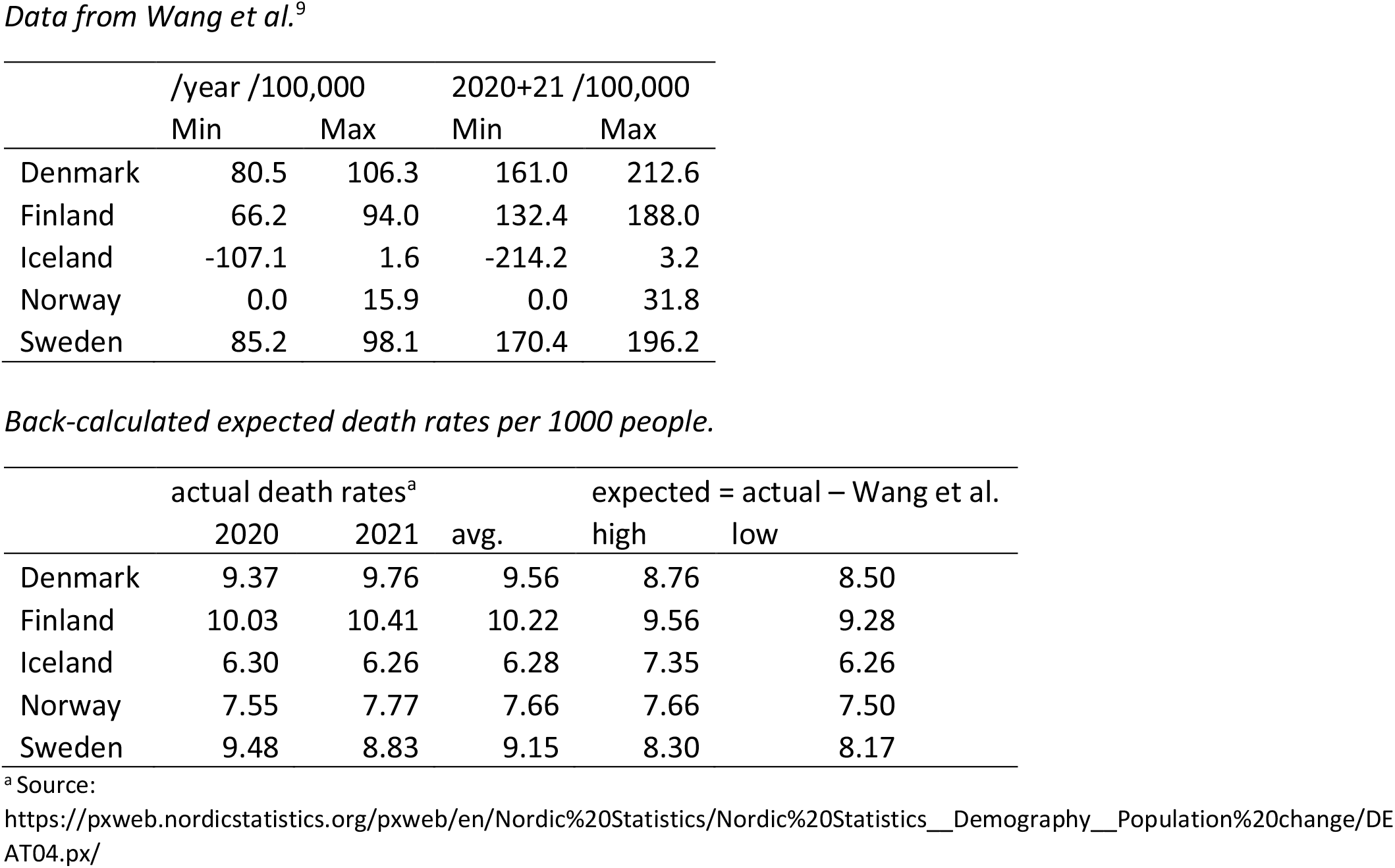
Back-calculated expected death rates from Wang et al. (average of 2020 and 2021).

**Table S3.**
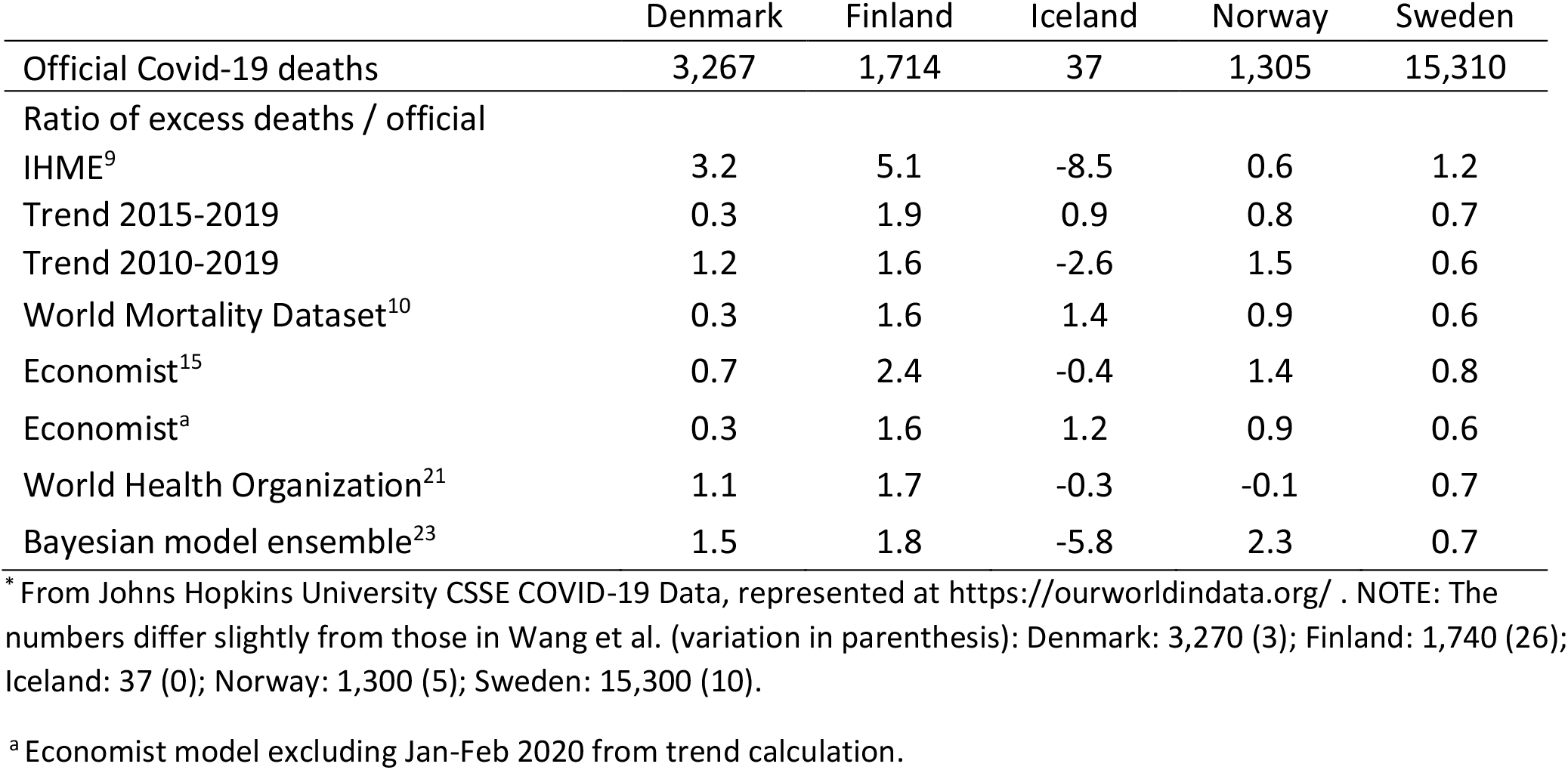
Ratio of excess deaths to official covid-19 deaths January 1, 2020 to December 31, 2021.*

**Table S4.** Total mean annual populations of the five Nordic countries 2010-2021.

|  | Denmark | Finland | Iceland | Norway | Sweden |
| --- | --- | --- | --- | --- | --- |
| **2010** | 5,547,683 | 5,363,352 | 318,041 | 4,889,252 | 9,378,126 |
| **2011** | 5,570,572 | 5,388,272 | 319,014 | 4,953,088 | 9,449,212 |
| **2012** | 5,591,572 | 5,413,970 | 320,716 | 5,018,572 | 9,519,374 |
| **2013** | 5,614,932 | 5,438,972 | 323,764 | 5,080,166 | 9,600,378 |
| **2014** | 5,643,475 | 5,461,512 | 327,386 | 5,137,429 | 9,696,110 |
| **2015** | 5,683,483 | 5,479,530 | 330,814 | 5,189,894 | 9,799,186 |
| **2016** | 5,728,010 | 5,495,302 | 335,439 | 5,236,151 | 9,923,085 |
| **2017** | 5,764,980 | 5,508,214 | 343,400 | 5,276,968 | 10,057,698 |
| **2018** | 5,793,636 | 5,515,524 | 352,720 | 5,311,916 | 10,175,214 |
| **2019** | 5,814,422 | 5,521,606 | 360,562 | 5,347,896 | 10,278,887 |
| **2020** | 5,831,404 | 5,529,542 | 366,463 | 5,379,474 | 10,353,442 |
| **2021** | 5,856,732 | 5,541,017 | 372,520 | 5,408,320 | 10,415,810 |
| **Change (%)**<br>**since 2010** | 5.6 | 3.3 | 17.1 | 10.6 | 11.1 |
| **2020 %**<br>**population**<br>**70+ years** | 14.5 | 16.1 | 9.9 | 12.6 | 14.9 |

Source:
https://pxweb.nordicstatistics.org/pxweb/en/Nordic%20Statistics/Nordic%20Statistics_Demography_Population%20size/POPU02.px/

**Table S5.**
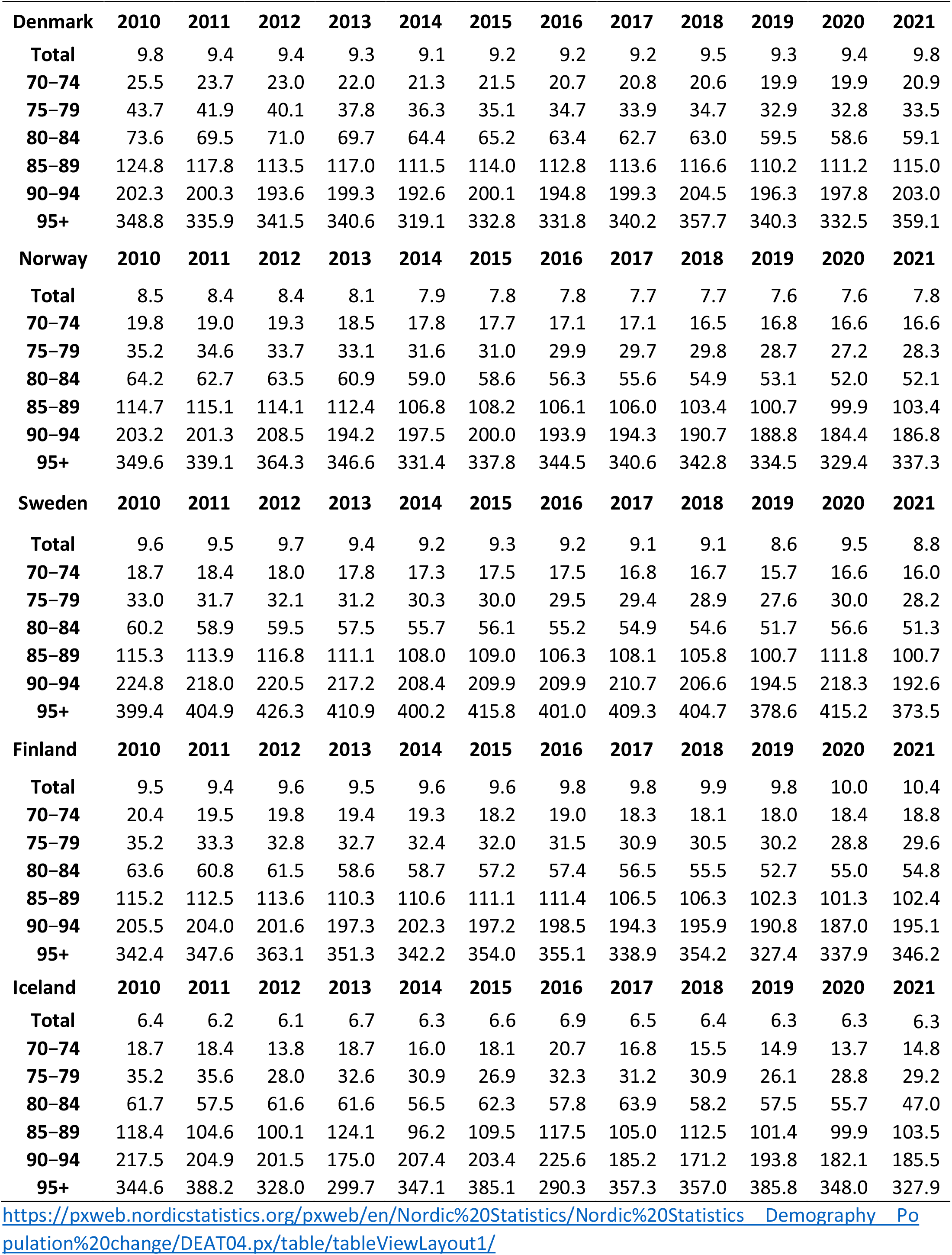
Age-specific death rates pr. 1000 people of Nordic countries 2020−2021.

**Table S6.** Estimated infection and infection fatality rates per November 14, 2021 by Barber et al.^12^ and corresponding estimates using the scale factors in Table 1 relative to Wang et al.^9^.

|  | Denmark | Finland | Iceland | Norway | Sweden |
| --- | --- | --- | --- | --- | --- |
| Predicted infection (%)<sup>12</sup> | 13.5 | 8.6 | 7.9 | 11.2 | 22.4 |
| Implied IFR (%)<sup>12</sup>: |  |  |  |  |  |
| IHME<sup>9</sup> | 1.2<br>(0.9–1.4) | 1.5<br>(1.2–1.8) | 0.1<br>(0.1–0.1) | 0.2<br>(0.1–0.3) | 0.7<br>(0.6–0.8) |
| 2015–19 trend | 0.12 | 0.55 | -0.01 | 0.30 | 0.40 |
| 2010–19 trend | 0.46 | 0.47 | 0.03 | 0.51 | 0.33 |
| WMD<sup>10</sup> | 0.11 | 0.45 | -0.02 | 0.30 | 0.38 |
| Economist<sup>15</sup> | 0.25 | 0.69 | 0.00 | 0.49 | 0.45 |
| Economist<sup>a</sup> | 0.12 | 0.46 | -0.01 | 0.31 | 0.38 |
| World Health Organization<sup>21</sup> | 0.43 | 0.49 | 0.00 | -0.03 | 0.44 |
| Bayesian model ensemble<sup>23</sup> | 0.55 | 0.52 | 0.07 | 0.81 | 0.39 |

<sup>a</sup> Economist model excluding Jan-Feb 2020 from trend calculation.

**Figure S1.**
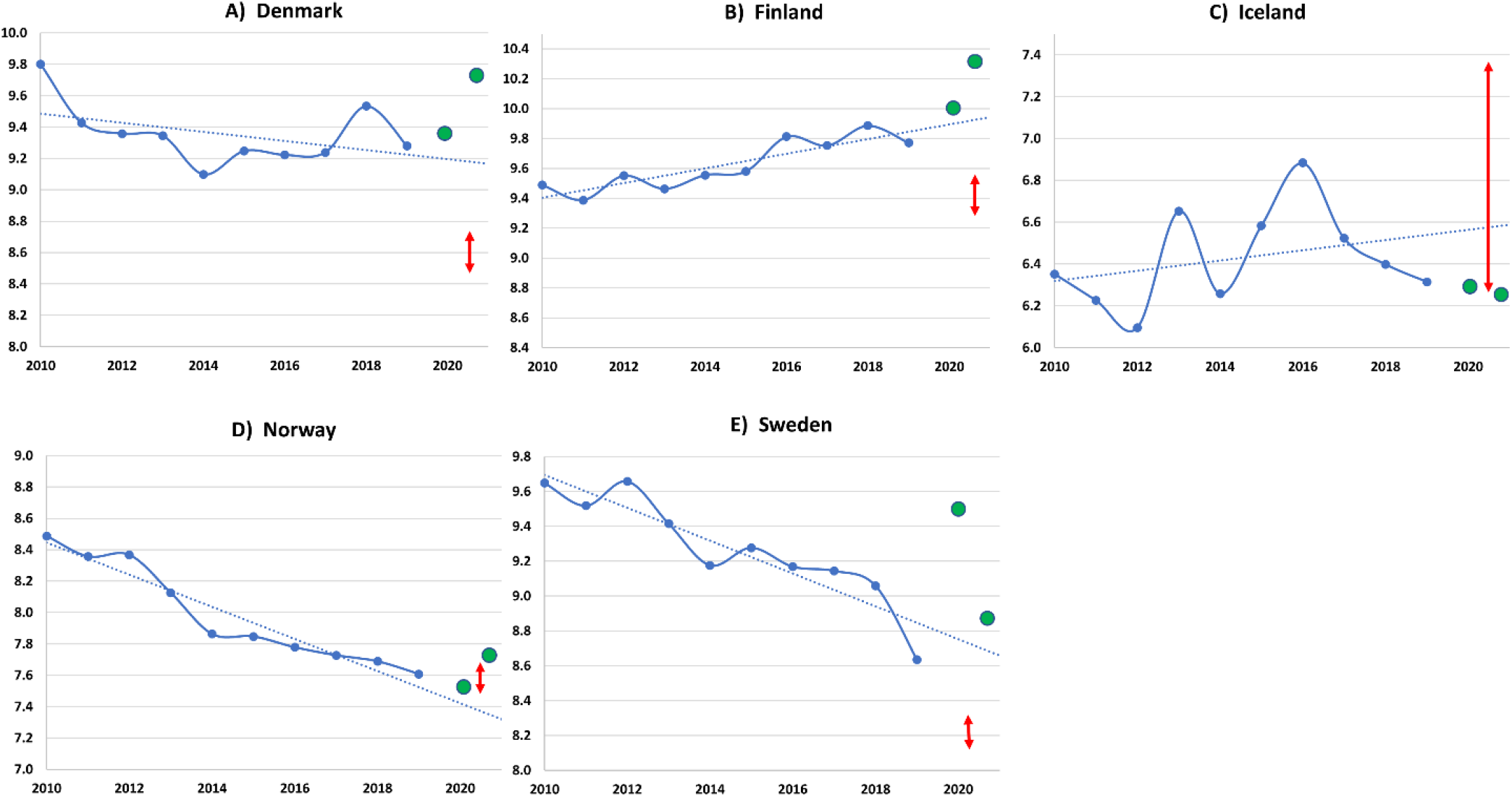
Linear regression trend lines for the annual actual all-cause mortality rates (deaths per 1000 people) of the Nordics (total for all age groups). The trends are compared with the actual death rates of 2020 and 2021 in green. The red lines indicate the approximate range of implied expected death rates estimated by recalculating excess death rates from Wang et al. (**A**) Denmark. (**B**) Finland. **(C)** Iceland. **(D)** Norway. **(E)** Sweden. Calculated from Table S2.

**Figure S2.**
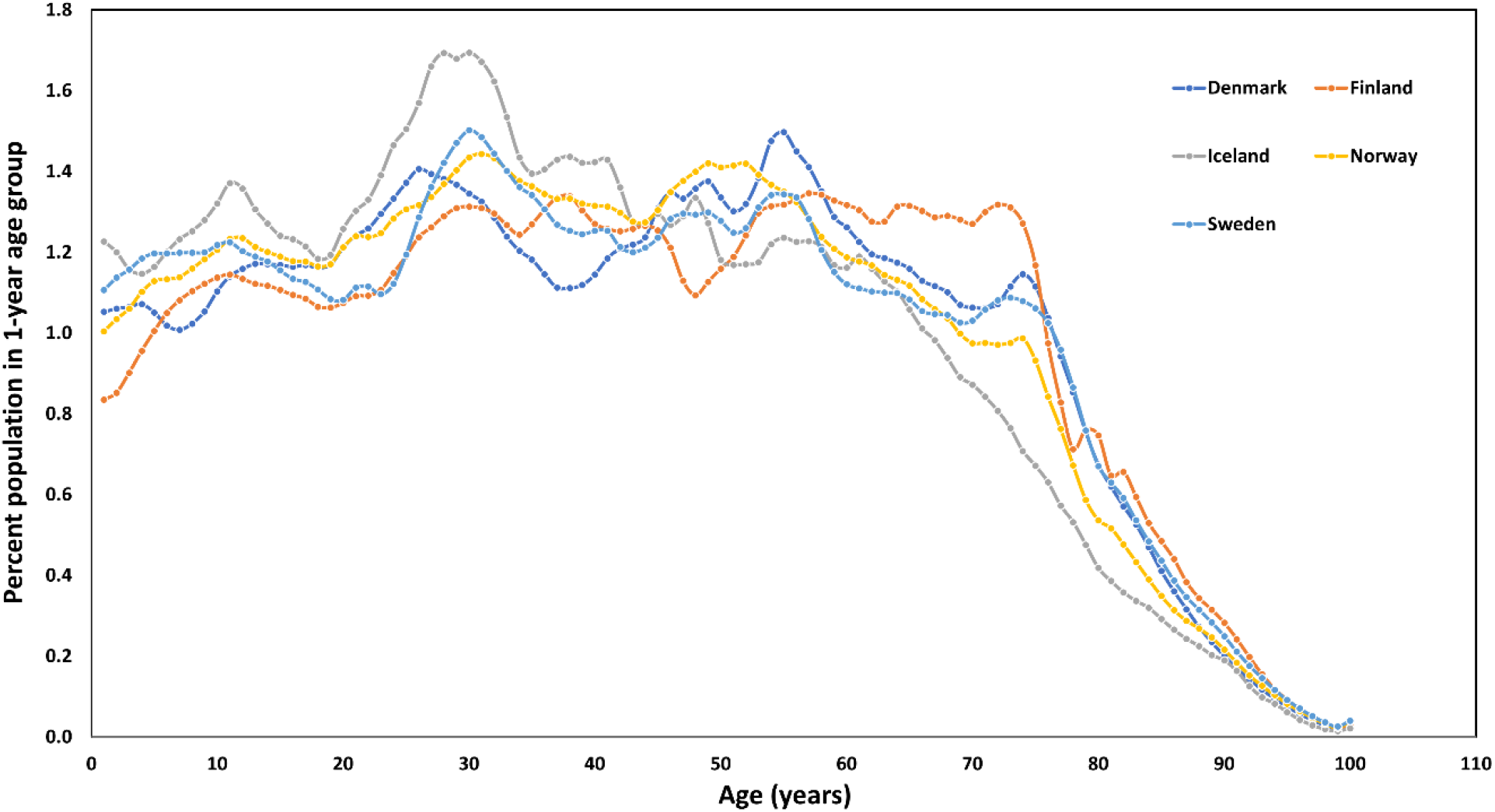
Mean population size per 1-year age group for the year 2020.

**Figure S3.**
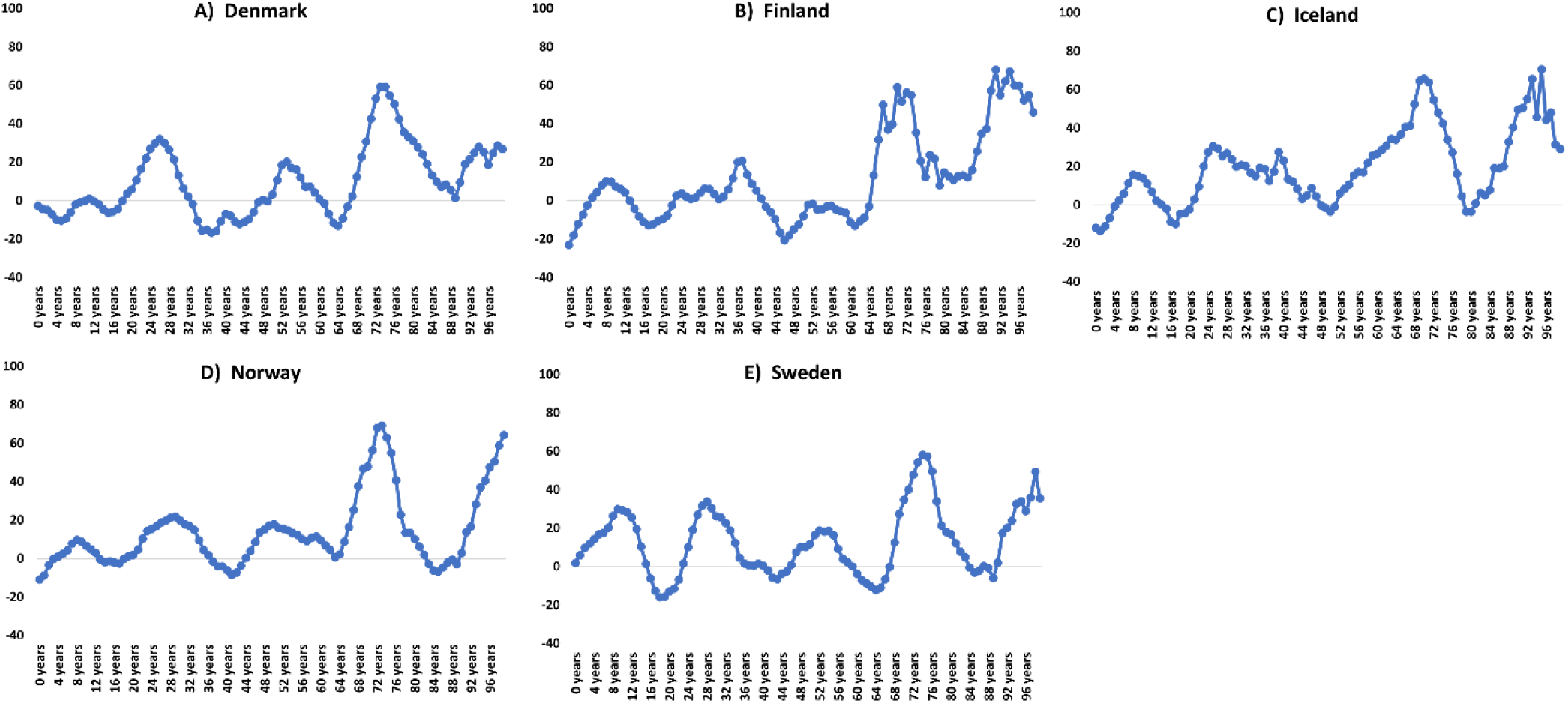
Change in absolute population (in %) of 1-year age groups for the Nordic countries 2010-2019. The last point represents 99 years and older. Source: https://pxweb.nordicstatistics.org/pxweb/en/Nordic%20Statistics/Nordic%20Statistics__Demography__Population%20size/POPU02.px/

**Figure S4.**
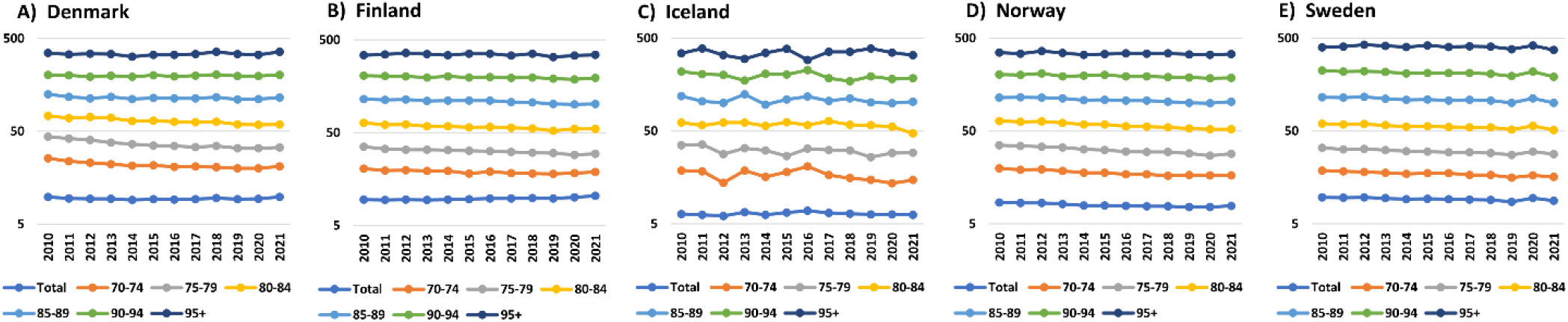
Logarithmic plot of Nordic age-specific death rates per 1000 people within five-year age groups. Based on final official deaths until 2021 per age group and mean annual population of each age group. (**A**) Denmark. (**B**) Finland. **(C)** Iceland. **(D)** Norway. **(E)** Sweden. The reverse mortality picture of Sweden in 2020 and 2021 is seen for several age groups.

**Figure S5.**
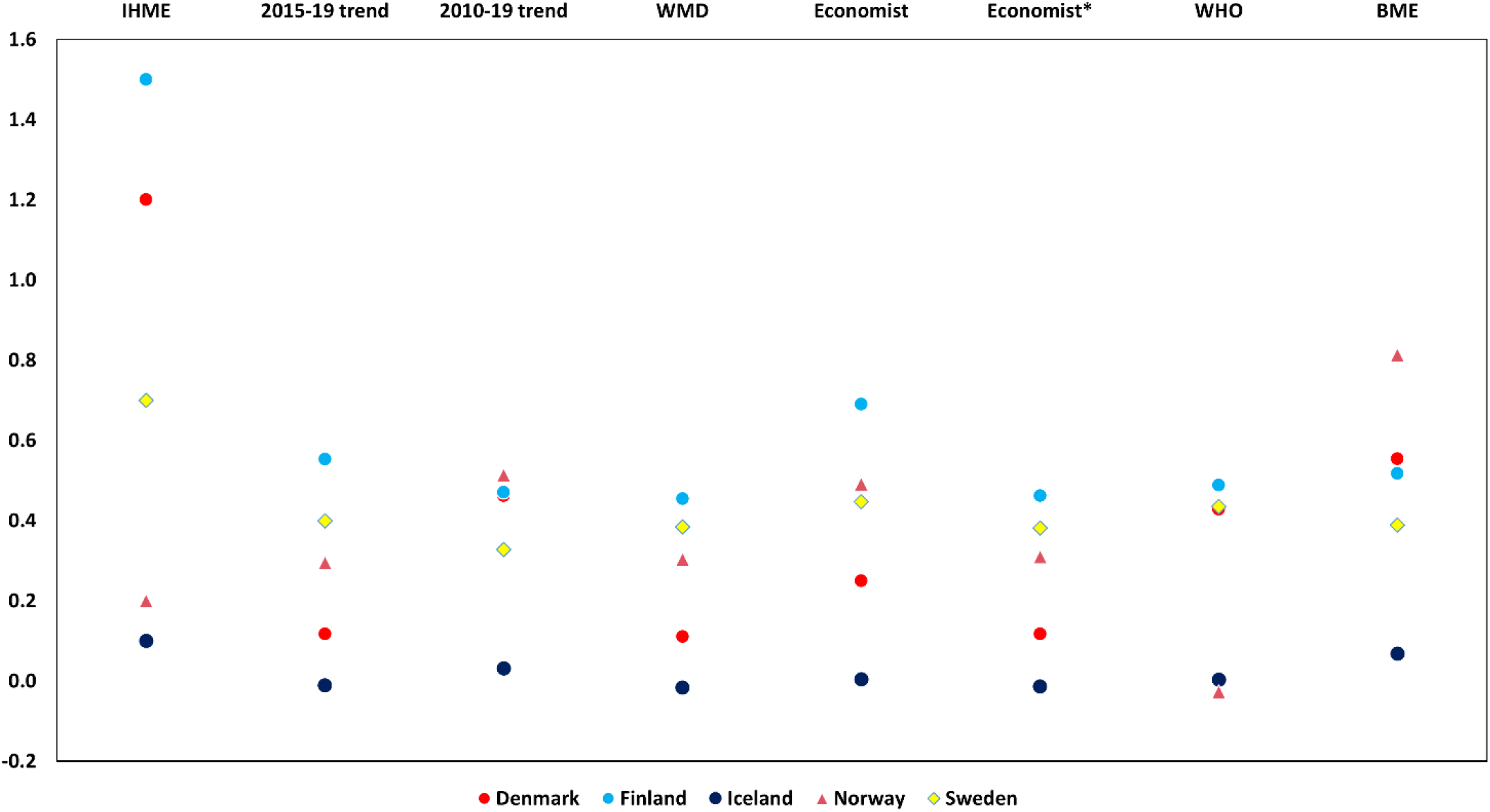
Infection fatality rates from IHME as reported in Barber et al.^12^ and corresponding estimates for other methods obtained using these IFR estimates multiplied by the scale factors of Table 1. (^*^ Economist model excluding Jan-Feb 2020 from trend calculation).

